# The Relationship Between COVID-19 Infection and Risk Perception, Knowledge, Attitude As Well As Four Non-pharmaceutical Interventions (NPIs) During the Late Period Of The COVID-19 Epidemic In China – An Online Cross-sectional Survey of 8158 Adults

**DOI:** 10.1101/2020.06.02.20120808

**Authors:** Hong Xu, Yong Gan, Daikun Zheng, Bo Wu, Xian Zhu, Chang Xu, Chenglu Liu, Zhou Tao, Yaoyue Hu, Min Chen, Mingjing Li, Zuxun Lu, Jack Chen

## Abstract

**Background:** So far, there has been no published population study on the relationship between COVID-19 infection and public’s risk perception, information source, knowledge, attitude and four non-pharmaceutical interventions(NPI: hand washing, proper coughing habits, social distancing and mask wearing) during the COVID-19 outbreak in China.

**Methods:** An online survey of 8158 Chinese adults between 22 February to 5 March 2020 was conducted. Bivariate associations between categorical variables were examined using Fisher exact test. We also explored the determinants of four NPIs as well as their association with COVID-19 infection using logistic regression.

**Results:** Of 8158 adults included, 57 (0.73%) were infected with COVID-19. The overwhelming majority of respondents showed a positive attitude (99.2%), positive risk perception (99.9%) and high knowledge levels that were among the strongest predictors of four highly adopted NPIs (hand washing:96.8%; proper coughing: 93.1%; social distancing:87.1%; mask wearing:97.9%). There was an increased risk of COVID-19 infection for those who not washing hands (2.28% vs 0.65%; RR=3.53: 95%CI: 1.53-8.15; P<0.009); not practicing proper coughing (1.79% vs 0.73%; RR=2.44: 95%CI: 1.15-5.15;P=0.026); not practicing social distancing (1.52% vs 0.58%; RR=2.63:95%CI:1.48 – 4.67; P=0.002); and not wearing a mask (7.41% vs 0.6%; RR=12.38:95%CI:5.81-26.36; P<0.001). For those who did practice all other three NPIs, wearing mask was associated with significantly reduced risk of infection compared to those who did not wear a mask (0.6% vs 16.7%; p=0.035). Similarly, for those who did not practice all or part of the other three NPIs, wearing mask was also associated with significantly reduced risk of infection. In a penalised logistic regression model including all four NPIs, wearing a mask was the only significant predictor of COVID-19 infection among four NPIs (OR=7.20; 95%CI:2.24-23.11; p<0.001).

**Conclusions:** We found high levels of risk perception, positive attitude, desirable knowledge as well as a high level of adopting four NPIs. The relevant knowledge, risk perception and attitude were strong predictors of adapting the four NPIs. Mask wearing, among four personal NPIs, was the most effective protective measure against COVID-19 infection with added preventive effect among those who practised all or part of the other three NPIs.

## Introduction

The unprecedented coronavirus disease of 2019 (COVID-19) global pandemic^1^ has changed the way our society operates. The total confirmed cases and deaths globally increased at an alarming rate^2^. The availability of an effective vaccine may still be many months away ^3 4^ and there is also no consensus on the use of antiviral drugs and other therapeutic agents^5 6^. Meanwhile, among the best hope for reducing mortality is societal preventative measures and providing timely and optimal critical care. As the list of countries in the grip of rapid spread of the COVID-19 is growing, many countries are or will be at the brink of further overwhelmed health care systems. Many countries have strengthened their non-pharmaceutical interventions (NPIs) in order to flatten the curve and reduce casualties ^7^ For the NPIs to be effective, one of the critical conditions is the public’s active participation and compliance. Since the lockdown of Wuhan city on 23^rd^ Jan 2019, China is the first country to have introduced NPIs with strict measures such as lockdown of cities and counties; compulsory mask wearing; isolation of suspicious cases; screening and contact tracing; quarantining people from high risk area for 14 days; as well as promoting hand washing, proper coughing habits, social distancing and self-isolation. However, there was no published evidence on relationships between COVID-19 infection, the Chinese public’s risk perception, information source, knowledge, attitude and personal NPIs during the middle towards the end of the epidemic.

Between 22 Feb 2020 to 5 March 2020 (the late period of COVID-19 epidemic in China)^2^, we conducted an online cross-sectional survey of Chinese residents to: 1) understand the risk perceptions, information source, knowledge, attitude and practice of Chinese public after the COVID-19 outbreak; 2) explore the determinants associated with the key personal NPIs (i.e., hand washing, proper coughing habits, social distancing, mask wearing); 3) estimate the risks between COVID-19 infection and the four NPIs; 4) understand potential risk compensating effects among four NPIs in relation to COVID-19 infection (e.g. can wearing mask further reduce risk of infection among those who with and without practicing other three NPIs?).

## Method

### Study sample

We conducted an online survey between 22 Feb 2020 (with total confirmed cases of 77k and daily cases of 1.5k) to 5 March 2020 (with a confirmed total case of 80.5k and daily cases of 151). Given that the whole of China was in lockdown during this period, it was almost impossible to conduct a random sample survey. We chose to conduct our study through the Chinese social media APP Wechat (similar to “WhatsApp”) and Weibo (similar to “Twitter”). We adopted a snow-balling sampling methodology through three social networks: 1) students and staff at Tongji Medical College and Chongqing Medical University; 2) Wanzhou District Centre for Disease Prevention and Control, Chongqing Municipality; 3) the study team. The inclusion criteria were: 1) the Chinese citizens who were currently living in Mainland China during the study period; 2) having a mobile phone or computer; 3) willing to answer all questions. The exclusion criteria were: 1) those who did not consent to participate; 2) those who did not answer all the questions; 3) the questionnaires completed in less than 2 minutes; 4) the repeated questionnaire from the same IP address. During the study period, the survey web page was browsed 21673 times with a total of 8431 questionnaires returned. After excluding those illegible questionnaires and those who were younger than 18 years old, the final study sample for the current study were 8158.

The Ethics Committee of Chongqing Medical University approved our study protocol. There was an introduction document before the study questionnaire that provided the respondents with the background, aims and estimated time (10 minutes) for completing the survey. Respondents were asked for their agreement to participate the study and to answer the questions faithfully and assured confidentiality and anonymity, and no individual data will be disclosed. After the confirmation of their willingness to participate the study voluntarily, the participants were directed to complete the online questionnaire. We plan to disseminate the results to study participants whenever appropriate.

**The roles of funding body:** The funding bodies played no roles in the study design, conduct, analysis, interpretation and the decision of the publication of the results.

### Measures

The study team developed the survey questionnaire through literature review, group discussion involving public who were not part of the research team and piloting. The final questions included in the survey were: 1) personal and family demographics: age, gender, location of residence, education, occupation, family monthly income, ever smoked during the last month (with over 100 cigarettes smoked over the lifetime), drinking alcohol during the last month, height and weight, if had been infected with COVID-19, marital status, if one of the family members is health professionals, the severity of the community infection where the respondent was living, if one of family members being part of local community efforts against COVID-19; 2) Perceived risk, attitude, information source, knowledge and four NPIs and if the respondent had repeatedly used a mask; 3)Self-isolation: if the respondents had a Chinese New Year party (2 days: 24 Jan – 25th Jan 2020) with invited guests, the main reason for the longest home stay family member; the main reason for the most often went out family member; the approach taken by the respondent when went out (i.e., shorten the time to avoid infection/as usual/ stay longer given the restrictions/uncertain).

### Statistical analysis

Frequencies of demographic, perceived risk, knowledge, attitude and four NPIs as well as self-isolation behaviours were described. The risks between COVID-19 infection and four binary NPIs were tested using Fisher exact tests. The absolute risk difference(RD), risk ratio (RR) and their 95% confidence intervals(CI) were also presented. We modelled the four NPIs using logistic regression based on theoretical framework of The theory of Reasoned Action (TRA) and Theory of Planned Behaviour (TPB) as developed by Ajzen and Fishbein^8^ which take into account the individual’s attitude and social norms as well as the individual’s perceived control as accurate predictors of behavioural intentions. The predictors included the demographic characteristics, social economics status, family and social environment, perceived risk of situation, attitude (belief) and respective knowledge on the four NPIs. We explored the risk between COVID-19 infection and four NPIs using similar approach but excluded distal determinates (i.e., knowledge, attitude and risk perception) of four NPIs based on a penalised maximum likelihood function logistic regression ^9 10^ which provides consistent estimates in situations of sparse event and total separation. The modelling results for four NPIs separately and combined (Model 1-Model 5) were compared to the results of the baseline model with only social demographic variables (Model 6). Risk ratio, odds ratio (OR) and their 95% CI were presented whereas appropriate. We explored the potential risk compensating effects among four NPIs through a pairwise NPIs comparison of infection rates and through the comparison of infection rates of wearing mask across a combination of other three NPIs. A flowchart of different sample sizes for the modelling of four NPIs and COVID-19 infections was presented (Figure 1). The data management and statistical analysis were done through SPSS v25 and Stata^TM^ v16. P value of 0.05 was considered as indicative of significance.

**Figure 1.**
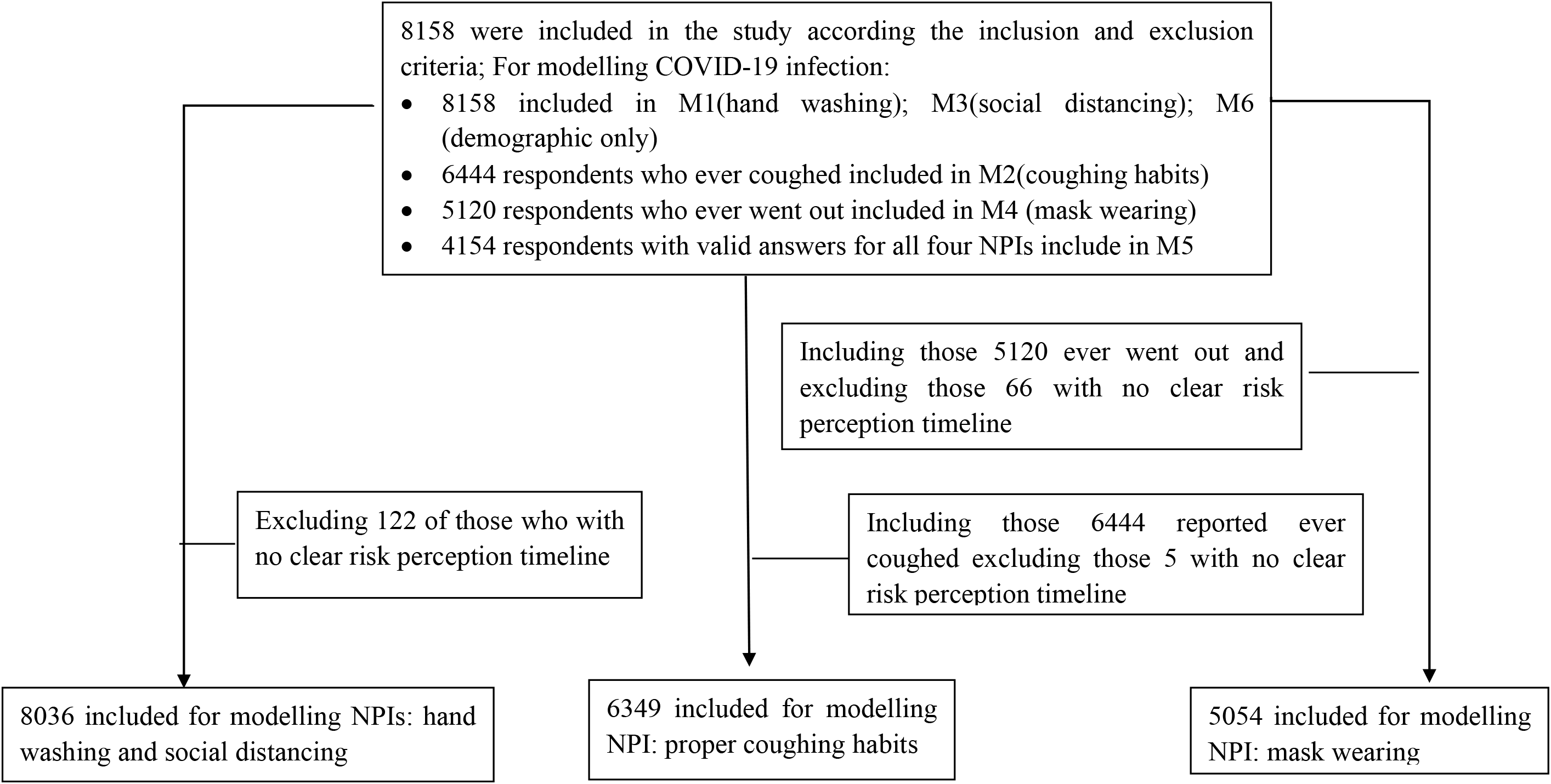
A flowchart of sample sizes for the modelling of 4 NPIs (Appendix: eTable 1) and COVID-19 infection (Appendix: eTable 3: Model 1 – Model 6)

## Results

### 1. Infection rate of COVID-19 and social demographic of respondents

In total, 8158 adults were included in the study and 57(0.7%) infected with COVID-19. The respondents were predominantly female (63%), younger age groups (18-39 years old: 62%), living in city (71.5%). Close to 45% of respondents had undergraduate and above education and close to one-quarter were students (Table 1). The last month family income had a large range (0-4,000,000 RMB) with close to 20% of them less than 2000RMB (approximately US$286 using exchange rate of $1=7RMB). Around 13% of respondents smoked and drank alcohol during the last month. Over half of them had normal Body Mass Index(BMI)^11^. Close to 72% of respondents had a partner and 54% were living with one. Close to 37% had a family member who were health professionals and 35% had a family member who was part of the local community efforts against COVID-19. Over 54% were from the area outside Hubei province with fewer than 100 cases, 42% from the areas outside Hubei province with more than 100 infected cases, 4% form Hubei province, the epicentre of Chinese COVID-19 epidemic (Table 1).

**Table 1.** The demographic characteristics of the study sample.

| Characteristics | N=8158 | % |
| --- | --- | --- |
| Age groups |  |  |
| 18-39 years | 5017 | 61.5% |
| 40-59 years | 2902 | 35.6% |
| >=60 years | 239 | 2.9% |
| Male (vs female) | 3020 | 37.0% |
| Currently living in City (vs rural area) | 5833 | 71.5% |
| Education |  |  |
| Primary School | 130 | 1.6% |
| High School | 2040 | 25.0% |
| Professional college | 2331 | 28.6% |
| University/Postgraduate | 3657 | 44.8% |
| Occupation |  |  |
| Health Professionals | 1373 | 16.8% |
| Government payee | 1814 | 22.2% |
| Factory workers/Managers | 1485 | 18.2% |
| Farmers | 313 | 3.8% |
| Students | 2006 | 24.6% |
| Others | 1167 | 14.3% |
| Family monthly Income |  |  |
| 0/1000 RMB | 607 | 7.4% |
| 1001/2000 RMB | 994 | 12.2% |
| 2001/4000 RMB | 1995 | 24.5% |
| 4001/6000 RMB | 1698 | 20.8% |
| 6001/8000 RMB | 714 | 8.8% |
| 8001/10000 RMB | 977 | 12.0% |
| 10001/20000 RMB | 620 | 7.6% |
| 20001/4000000 RMB | 216 | 2.6% |
| Not sure/unanswered | 337 | 4.1% |
| Ever smoked during the last month* (yes) | 1087 | 13.3% |
| Drinking during the last month |  |  |
| Yes | 1088 | 13.3% |
| Give up | 308 | 3.8% |
| Don't drink | 6762 | 82.9% |
| Body Mass Index |  |  |
| underweight | 1574 | 19.3% |
| Normal | 4263 | 52.3% |
| overweight | 1264 | 15.5% |
| Obese | 133 | 1.6% |
| Not available | 924 | 11.3% |
| Infected with COVID-19?(Yes) | 57 | 0.7% |
| Do you currently live with your partner? |  |  |
| Yes | 4420 | 54.2% |
| No | 1434 | 17.6% |
| I don't have a partner | 2304 | 28.2% |
| Do you have a health professional family member? (yes) | 3001 | 36.8% |
| From the area with community infection |  |  |
| Hubei Province | 319 | 3.9% |
| Outside Hubei province with 100+ cases | 3400 | 41.7% |
| Other | 4439 | 54.4% |
| Having a family member being part of local community efforts against COVID-19 | 2835 | 35.1% |
| Total | 8158 | 100.0% |

\*Ever smoked was defined as with 100+ cigarettes smoked over the lifetime.

### 2. Risk perception, information source, knowledge, attitude, four NPIs and self-isolation

Close to 7% of respondents were aware of the seriousness of the situation on 11 Jan 2020 when the first COVID-19 related patient death was announced by the Wuhan Health Commission; 39% on 20 Jan 2020 with the announcement of COVID-19 transmission among humans; 29% on 23^rd^ Jan 2020 with the lockdown of Wuhan city; 24% after 24^th^ Jan 2020 due to the activation of Level 1 pubic emergency responding scheme by local governments and strict measures and lockdown of neighbourhood or villages.^12^ Nine respondents (0.1%) did not think it was serious at the time (Figure 2). Overwhelmingly, the majority (99.2%) strongly agreed with the position that the fight against COVID-19 is everyone’s responsibility (Table 2). Close to 97% perceived governmental websites, APP and public media as the most authoritative source of information; 90% felt that governmental websites, APP and public media were also the most involved source of information; 99.6% of respondents knew why and how to wash hands properly during the COVID-19 outbreak period; 97.2% were aware of the proper procedures when coughing (turning away from people and covering mouth and nose when coughing and washing hands afterwards); 97.8% knew the right way of practicing social distancing (i.e., keeping social distance more than 1 meter and avoiding close contact with those who had fever or cough); and 99.9% knew why and how to wear a mask. The overwhelming majority also reported that they translated this knowledge into practice: 96.8% when washing hands; 93.1% when coughing; 87.1% when social distancing; and 97.9% when wearing a mask (Table 2).

**Table 2.** Attitude, source of information, knowledge and four NPIs.

|  | N=8158 | % |
| --- | --- | --- |
| Agree with that the fight with COVID-19 is everyone's job (Yes) | 8094 | 99.2% |
| Perceived most authoritative source of information |  |  |
| Governmental websites/APP/Public Media | 7902 | 96.9% |
| Weibo/Webchat friends | 128 | 1.6% |
| QQ/webchat groups | 64 | 0.8% |
| Family/Friends | 40 | 0.5% |
| Other | 24 | 0.3% |
| Perceived most interested source of information |  |  |
| Governmental websites/APP/Public Media | 7396 | 90.7% |
| Weibo/Webchat friends | 470 | 5.8% |
| QQ/webchat groups | 218 | 2.7% |
| Family/Friends | 60 | 0.7% |
| Other | 14 | 0.2% |
| Know why and how to wash hands (Yes) | 8129 | 99.6% |
| Know the proper habit when coughing (Yes) | 7927 | 97.2% |
| Know why and how to practice social distancing (Yes) | 7975 | 97.8% |
| Know why and how to wear a mask in public (yes) | 8146 | 99.9% |
| Washing hands (Yes) | 7895 | 96.8% |
| Acting in proper habit when coughing (Yes) * | 5997 | 93.1% |
| Practicing social distancing (Yes) | 7104 | 87.1% |
| Wearing a mask when went out (yes)<sup>§</sup> | 5012 | 97.9% |
| Repeated use of a mask (yes)<sup>§</sup>? | 5117 | 99.9% |

\*: Only 6444 included as 1714 person reported that they did not cough during the last month.
§: Only 5120 respondents of who went out during the period after outbreak included

**Figure 2.**
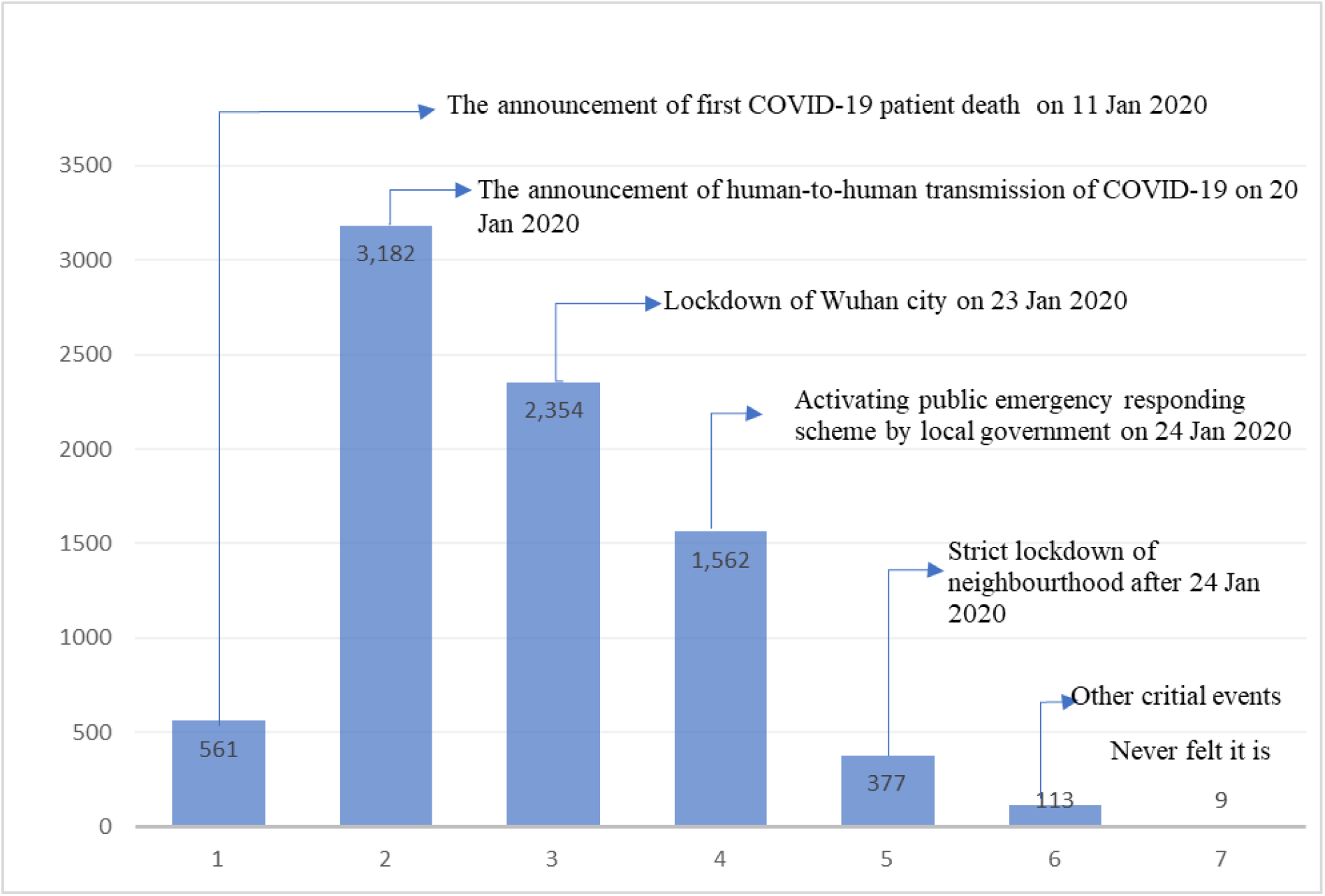
When felt the situation was very serious

Approximately 23% of respondents reported had a Chinese near year party with invited guests (Appendix; eTable 2). The major reasons for the family member who stayed longest at home included: 1) comply with the call from government (65.6%); 2) self/compulsory isolation (13.6%); 3) fear of the virus (5.7%); 4) the focus of family protection (3.8%); and 5) no mask (3.0%). The main reason for going out were: 1) shopping(40.5%); 2) partaking in work related to controlling COVID-19(32.1%); 3) doing usual job (21.4%); 4)having a walk (2.6%); 5) getting delivery(0.7%); and 6) socializing/dinner party (0.3%). Over 74% shortened the time to avoid infection when they went out; close to 20% acted in the usual way; and 1% (50) stayed longer than usual, given the restrictions and difficulties to be able to get out home (Appendix 1; eTable 2).

### 3. The determinants of four NPIs

Our multivariate models found that the knowledge of the relevant NPIs was one of the strongest predictors of that behaviour (OR=22.6 for hand washing, OR=4.26 for social distancing, all Ps < 0.001, and positive knowledge level associated with proper coughing habit and mask wearing were 100% and excluded from the models, Appendix: eTable 1).

The belief that task of fighting against COVID-19 is everyone’s responsibility was positively associated with the hand washing (OR=5.59;P<0.001), social distancing (OR=3.76; p<0.001) and mask wearing(OR=26.89; p<0.001). Those who perceived the seriousness of the situation before the lockdown of Wuhan city were more likely to practice hand washing (OR=1.43; p<0.05), proper coughing habit (OR=1.54; p<0.05). Those who had a family member being of the local community efforts against COVID-19 and people from outside Hubei province were more likely associated with positive NPIs. In comparison to those who currently living with a partner, those who did not have partner were less likely to practice hand washing (OR=0.57; p<0.05), proper coughing habits (OR=0.59; p<0.05), but were more likely to practice social distancing (OR=1.64; p<0.001). The respondents who have a normal body weight were more likely to practicing social distancing (OR=1.21; p<0.05) than those who were underweight. Those non-smokers were more likely to practice social distancing (OR=1.38; p<0.01) than smokers. Family income, education, occupation, living area, sex and age groups demonstrated differential impacts on different NPIs (Appendix 1; eTable 1).

### 4. Risk association between COVID-19 infection and hand washing, coughing habits, social distancing and mask wearing

The bivariate analyses between individual NPI and COVID-19 infection showed that there was a significantly increased risk of COVID-19 infection (Table 3) for those who did not wash hands (2.28% vs 0.65%: RR=3.53; 95%CI: 1.53-8.15; P<0.009); who did not practice proper coughing (1.79% vs 0.73%: RR=2.44; 95%CI:(1.15-5.15);P=0.026); who did not practice social distancing (1.52% vs 0.58%; RR=2.63:95%CI:(1.48 - 4.67); P=0.002); and who did not wear a mask (7.41% vs 0.6%; RR=12.38:95%CI:(5.81-26.36); P<0.001). The adjusted ORs were 4.68(95%CI: 1.78-12.28) for not washing hands; 2.79(95%CI:1.19-6.57) for not practicing proper coughing; 2.14(95%CI: 1.17-3.94) for not practicing social distancing and 12.76 (95%CI:5.00-32.57) for not wearing a mask. The model which adjusted all four NPIs plus social demographic variables (Model 5; Appendix 1; eTable3:M5) showed that not wearing a mask was the only significant predictor of infection (OR=7.20; 95%CI:2.24-23.11;). In comparison to those who had only primary school education, those with high school qualification were less likely to be infected(OR=0.12; p<0.001). This was similar to those who had professional college qualifications (OR=0.10; p<0.001) or with university degrees (OR=0.15; p<0.001; Appendix 1: eTable 3: Model 6). Non-smokers were less likely to be infected than smokers (OR=0.40; p<0.01) and those who with a monthly family income of 8001-10000RMB were less likely to be infected than those who having a monthly family income less than 1000RMB (OR=0.20; p<0/05) (Appendix 1:eTable 3:M6).

**Table 3.** The association between COVID-19 infection and four NPIs with adjusted OR and 95%CI from penalised logistic regression models.

| Risk of COVID-19 infection (57/8158: 0.70%) |  |  |  |  |  |  |
| --- | --- | --- | --- | --- | --- | --- |
| Risk |  | Risk Difference(95%CI) | Risk Ratio (RR) | P | Adjusted OR (95%CI)<sup>§</sup> | Adjusted OR (95%CI)<sup>#</sup> |
| Washing hands (n=8158) |  |  |  |  |  |  |
| No | (6/263) 2.28% | 1.63% (-0.18% - 3.45%) | 3.53(1.53 - 8.15) | 0.009 | 4.67**(1.86 - 11.74) | 1.82 (0.40 - 8.32) |
| Yes | (51/7895) 0.65% |  |  |  |  |  |
| Acting properly when coughing (n=6444) |  |  |  |  |  |  |
| No | (8/447) 1.79% | 1.06% (-0.19% - 2.3%) | 2.44(1.15 - 5.15) | 0.026 | 2.78* (1.22 - 6.33) | 1.88 (0.60 - 5.94) |
| Yes | (44/5997) 0.73% |  |  |  |  |  |
| Practicing social distancing (n=8158) |  |  |  |  |  |  |
| No | (16/1054 )1.52% | 0.94% (0.18% - 1.7%) | 2.63(1.48 - 4.67) | 0.002 | 2.13* (1.17 - 3.85) | 1.07 (0.46 - 2.46) |
| Yes | (41/7104) 0.58% |  |  |  |  |  |
| Wearing a mask when went out (n=5120) |  |  |  |  |  |  |
| No | (8/108) 7.41% | 6.81% (1.87% - 11.75%) | 12.38(5.81 - 26.36) | <0.001 | 11.03*** (4.53 - 26.84)) | 7.20*** (2.24 - 23.11) |
| Yes | (30/5012) 0.60% |  |  |  |  |  |

\*\*\* p<0.001, \*\* p<0.01, \* p<0.05; §: Results from M1-M4 logistic regression model adjusting for social demographic variables plus washing hands, acting properly when coughing, practicing social distancing, wearing a mask when went out, respectively.(Appendix 1; eTable2: M1-M4); #: Results from M5 logistic regression model adjusting for social demographic variables plus all four behaviours(Appendix 1;eTable2; M5).

### 5. Potential risk compensating effects among four NPIs against COVID-19 infection

The pairwise distributions of COVID-19 infection rate among four NPIs are presented (Table 4). Wearing mask (versus not) was associated with significantly reduced risk of COVID-19 infection among those who practiced hand washing (0.6% vs 5.3%; RR=0.11; p<0.001), proper coughing (0.7% vs 3.9%; RR=0.18; p=0.019) and social distancing (0.5% vs 16.7%; RR=0.03; P=0.002). Hand washing showed a trend towards further reduced risk of infection for those who did not practice social distancing (RR=0.25; p=0.053). Among those who did not practice social distancing, persons who had proper coughing habits were associated with a reduced risk of infection compared to those who did not have proper coughing habits (1.3% vs 4.4%; RR=0.29; p=0.048). The potential added protecting effects of mask wearing on different combinations of the other three NPIs are presented in Figure 3. For those who did practice all three NPIs (i.e, hand washing(H), proper coughing(C), social distancing (S): HCS), wearing a mask were associated with significantly reduced risk of infection compared to those who did not (0.6% vs 16.7%; p=0.035). Similarly, for those who did not practice all other three NPIs, wearing a mask was also associated with significantly reduced risk of infection compared to not wearing a mask (Figure 3).

**Table 4.** The COVID-19 infection rates (%) and potential pairwise risk compensating effect among four NPIs with relative risk (RR) and P values from exact tests.

|  |  | Mask wearing |  |  |  |  |  | Hand washing |  |  |  |  |  | Proper coughing |  |  |  |  |  |
| --- | --- | --- | --- | --- | --- | --- | --- | --- | --- | --- | --- | --- | --- | --- | --- | --- | --- | --- | --- |
|  |  | no |  | yes |  | RR (95%CI) | P | no |  | yes |  | RR (95%CI) | P | no |  | yes |  | RR (95%CI) | P |
|  |  | N | % | N | % |  |  | N | % | N | % |  |  | N | % | N | % |  |  |
| **Hand washing** | No | 13 | 23.1 | 147 | .7 | 0.03(0.003-0.26) | 0.002 | - | - | - | - |  |  | - | - | - | - |  |  |
|  | Yes | 95 | 5.3 | 4865 | .6 | 0.11 (0.04-0.29) | <0.001 | - | - | - | - |  |  | - | - | - | - |  |  |
| **Proper coughing** | No | 11 | 36.4 | 318 | .6 | 0.02(0.004-0.08) | <0.001 | 74 | 4.1 | 373 | 1.3 | 0.33 (0.08-1.35) | 0.131 | - | - | - | - |  |  |
|  | Yes | 76 | 3.9 | 3749 | .7 | 0.18 (0.05-0.57) | 0.019 | 118 | 1.7 | 5879 | .7 | 0.42 (0.10-1.72) | 0.214 | - | - | - | - |  |  |
| **Social distancing** | No | 96 | 6.3 | 958 | 1.0 | 0.17 (0.06-0.45) | 0.002 | 58 | 5.2 | 996 | 1.3 | 0.25 (0.73-0.86) | 0.053 | 90 | 4.4 | 776 | 1.3 | 0.29(0.09-0.91) | 0.048 |
|  | Yes | 12 | 16.7 | 4054 | .5 | 0.03 (0.01-0.11) | 0.002 | 205 | 1.5 | 6899 | .6 | 0.38 (0.12-1.21) | 0.114 | 357 | 1.1 | 5221 | .7 | 0.58 (0.21-1.63) | 0.304 |

**Figure 3.**
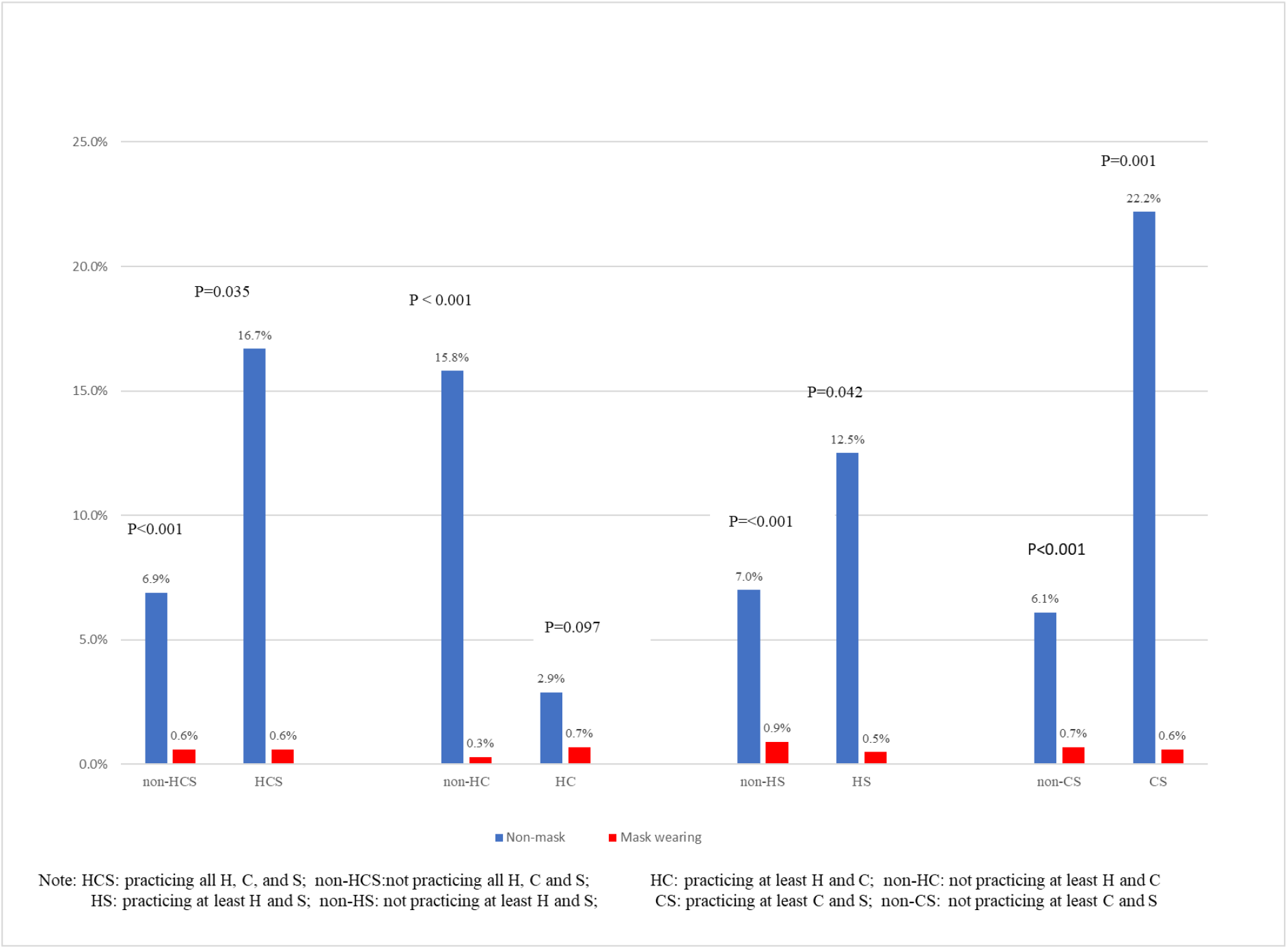
The COVID-19 infection rates between mask wearing and the combinations of NPIs: hand washing (H), proper coughing habit (C), soical distancing (S)

## Discussion

To our best knowledge, our study is the first comprehensive report of the COVID-19 infection rate, perceived risk, knowledge, attitude, four NPIs and self-isolation of a national-wide adult sample amidst the late period of the COVID-19 epidemic in China. We found that most respondents were aware of the seriousness of the outbreak at different time periods and believed that it is everyone’s responsibility to fight against the spread of COVID-19. The positive attitude, earlier risk perception and relevant knowledge were among the strongest predictors of hand washing, proper coughing habits, social distancing and mask wearing. Different social demographic factors also contributed to different NPIs. Those having only primary school education, with little family income, and being a smoker were associated with increased risk of COVID-19 infection. Mask wearing, among the four practices, was the most important protecting factor against COVID-19 infection with added preventive effect among those who practised all or part of the other three NPIs.

Our findings of high levels of knowledge among the Chinese public are consistent with the results of a previous study^13^. However, the previous online survey study was conducted at a much earlier stage (i.e., 27 Jan to 1 Feb, 2020) with a smaller sample and over half of respondents from Hubei province which did not include the knowledge and behaviours of hand washing, proper coughing habit, social distancing and self-isolation (but with mask wearing included). The widespread use of mobile phone, internet, the social media APP such as Wechat (with estimated 1.1 billion registered accounts in China in 2019)^14^ have significantly increased the speed and scope of information transmission in China and may be instrumental in the dissemination of COVID-19 related knowledge and information. This also helped by the fact that over 90% of respondents believed that the government web sites, public media and APP were the most authoritative and involved source of information for COVID-19. Our results suggested that respondents had an extremely positive attitude that fighting against COVID-19 is everyone’s responsibility and its association with positive NPIs highlighted the success of a national wide campaign in instilling the concept that everyone can and should make necessary contributions towards the fight against COVID-19.

Our study’s finding that the early perceived seriousness of the situation is a strong predicting factor for the use of NPIs reinforces the importance of transparency and timely dissemination of the critical information of COVID-19 pandemic. Our study showed that around 22% of respondents had a party during Chinese New Year (24 Jan – 25 Jan) which is the most important festive in the Chinese tradition. This was the period immediate after the lockdown of Wuhan city (on 23^rd^ Jan) with much confusion for those who lived outside Hubei province and there was no social distancing and self-isolation in place at the time. Such gatherings could be avoided if the public had been equipped with the real-time knowledge of the danger and seriousness of the situation. Evidence also provided support for the concept that the early swift NPIs by governments and education of public on the seriousness of situation are critical in slowing the spread and flattening the curve^15^. Our study demonstrated that family influences (in particular in those with health professionals and with someone being part of community team in fighting against COVID-19) could have a significant positive impact on individual’s behaviours. Respondents with various demographic characteristics had exhibited different NPIs. These findings may provide further opportunities for developing tailored health education campaigns and health policy interventions on the segments of the population in order to maximize the effects of the NPIs. For example, specific policies and education could target the smokers, and those younger population for encouraging certain behaviours (such as proper coughing habit) and older segment population on the other behaviours (such as self-isolation, social distancing and mask wearing).

Our study found almost universal acceptance of the importance of mask wearing and very high proportion (97.9%) wore a mask in public after the outbreak. Our study found that mask wearing, among four personal NPIs, was the most important protective measure against COVID-19 infection. This may have policy implications. The Chinese public were accepting towards the concept of mask wearing possibly due to factors such as the previous SARS epidemic experience^16^, coordinated national wide education campaign, the earlier recognition of existence of asymptomatic virus carriers, the strict measures in reinforcing such a role (e.g., in shopping centres, in public transportation, etc), the coordinated efforts in rationing the supply to families over the shortage period. The necessity in wearing mask in public may be controversial in different countries and agencies^17-21^ despite the positive evidence in favouring wearing a mask in a simulated environment^22 23^. It is likely to be an evolving policy option depending on several factors, including the availability of masks and fair distribution channels among the society. Our findings that mask wearing had added preventive effect even among those who did practice all or part of other three NPIs provided contradictive evidence towards the notion that other three NPIs alone are sufficient in preventing COVID-19 infection. It also did not support notion that wearing a mask could even increase the risk of infection through more facial contacts. For those countries still in the grip of pandemic or who are considering reopening their economy, a policy of encouraging or requiring public to wear a mask may have a positive impact especially in highly populated areas or in settings where other NPIs are very difficult to implement (such as in bus, airplane, or shopping centre). During the study period, there were still 3% of respondents who reported that ‘no mask’ was the main reason for stopping them from going out and most respondents had repeatedly used the same mask. Given the likelihood of surged demand for masks during an outbreak, public health agencies and related authorities may also need to provide practical and evidence-based guidance on when and how to appropriately reuse a mask. China contributed over half of the global mask manufacturing output before the outbreak but still faced the shortage of mask over the epidemic period^24^. It is important for governments and international agencies to rethink the adequacy of and better approaches towards their strategic stockpiles of masks and other personal protection equipment (PPE) for the current and future pandemics.

Our study has several strengths. First, it was the largest study of its kind to cover the most critical period of the COVID-19 outbreak in China. Second, our study design and analysis were driven by theory and policy needs and included many factors such as demographics, social economic status, family contextual factors, risk perception, knowledge, attitude and personal practices. Third, the adoption of the internet survey methodology enabled us to complete our study in a critical period and cost-effective manner. Our study also has several limitations. First, our study sample have disproportionately more female, well-educated and less smokers, reflecting a typically young and healthy cohort in similar surveys. Thus, the frequencies of desirable knowledge levels and health behaviours may be over-estimated while less desirable outcomes (such as lower family monthly income) may be under-estimated. However, the modelling results may be less susceptible to these potential biases. Second, our study results were from a particular period of the outbreak and most of the respondents were from outside Hubei province. The generalization of the results to other settings and countries may be limited. Third, our study was a cross-sectional population survey and the association found between the predictors and outcomes should be interpreted with caution and further intervention studies are needed in confirming our findings. Fourth, despite the relatively large sample size, the total cases of COVID-19 infection were still small so that the relationship between NPIs and the covid-19 infection should be confirmed by other larger epidemiological studies. Fifth, the potential risk compensating effects of mask wearing against other NPIs should be considered as being of a hypothesis-generating in nature given the potential limitations outlined above. Common to any observational studies with multiple outcomes and modelled with different effective sample sizes, the interpretations and generalisation of the results should be strictly limited to the same setting and be cautious with multiple tests risks.

## Conclusions

Our study found high level of risk perception, positive attitude, desirable knowledge and practices in hand washing, proper coughing habit, social distancing and mask wearing among a large cohort of Chinese adults. Our study also found that the relevant knowledge, risk perception and attitude were among the strongest predictors of the four NPI. Mask wearing, among the four NPIs, was the predominating protecting measure against COVID-19 infection with added preventive effect among those who practiced all or part of other three NPIs. Our findings of many different predictors on different personal NPIs may also provide the possibility for further tailored health policy interventions. The study also emphasises the importance at an international level of sharing information in a collaborative way in order to learn from everyone’s experiences about what interventions worked well and what was the impact of issues that may have resulted in poor outcomes such as delayed and misinformed actions.

## Data Availability

The datasets generated and/or analysed during the current study are not publicly available due ongoing research effort and data analysis by the team but are available from the corresponding author on reasonable request.

**List of Abbreviations**
BMI: Body Mass Index;
CI: Confidence Interval
COVID-19: Coronavirus disease of 2019
NIPs: non-pharmaceutical interventions;
OR: Odds ratio;
RD: Risk difference;
RR: Risk ratio;
TPB: Theory of Planned Behaviour;
TRA: Theory of Reasoned Action

## Ethics approval and consent to participate

The Ethics Committee of Chongqing Medical University approved our study protocol.

There was an introduction document before the study questionnaire that provided the respondents with the background, aims and estimated time (10 minutes) for completing the survey. Respondents were asked for their agreement to participate the study and to answer the questions faithfully and assured confidentiality and anonymity, and no individual data will be disclosed.

## Consent for publication

Not applicable.

## Authors’ contributions

Hong Xu(HX), Yong Gan(YG), Daikun Zheng(DKZ), Bo Wu(BW), Xian Zhu(XZ), Chang Xu(CX), Chenglu Liu(CLL), Zhou Tao(ZT), Yaoyue Hu(YYH), Min Chen(MC), Mingjing Li(MJL), Zuxun Lu1(ZXL), Jack Chen(JC)

HX, YG, ZXL conceived and designed the study. HX, YG, DKZ, BW, XZ, CX, CLL, ZT, MC, and MJL participated in the acquisition of data. JC, HX, XZ conceptualised the theoretical and analytical framework and conducted data management and statistical analysis. ZXL, YYH provided advice on the methodology. JC, HX, YG, XZ conducted literature review and HX, YG, and XZ provided the first draft of the manuscript. JC produced all final tables and figure and the final draft for review. ZXL, YYH and other authors revised the manuscript. All authors read and approved the final manuscript. JC is the guarantor of this work and has full access to all the data in the study and takes responsibility for its integrity and the accuracy of the data analysis.

## Acknowledgement

We would like to acknowledge the School of Public Health & Management of Chongqing Medical University and The Science and Technology Association of Chongqing Municipal Government for the financial and in-kind support of this research project.

## Appendix 1

**eTable 1.**
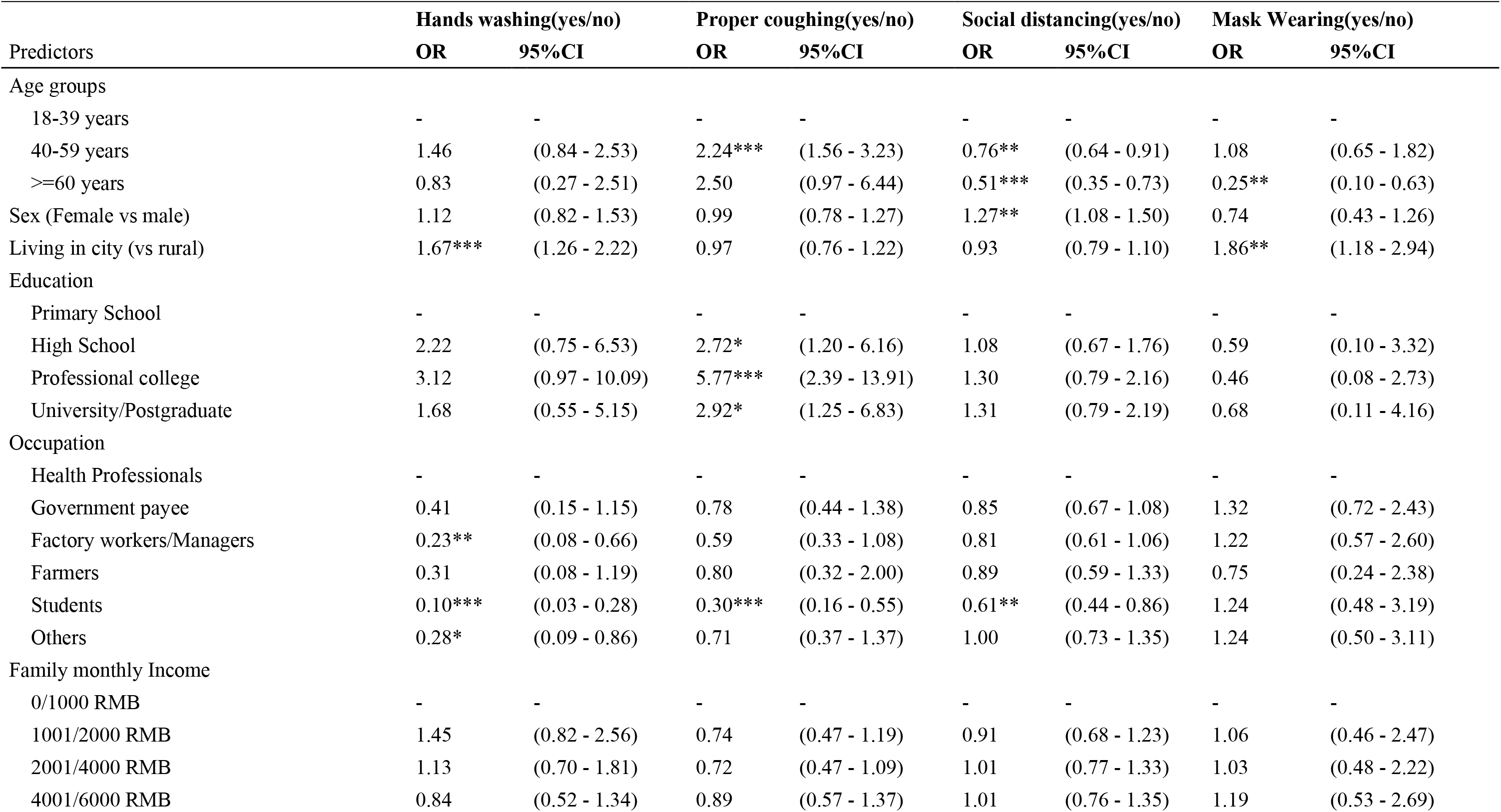
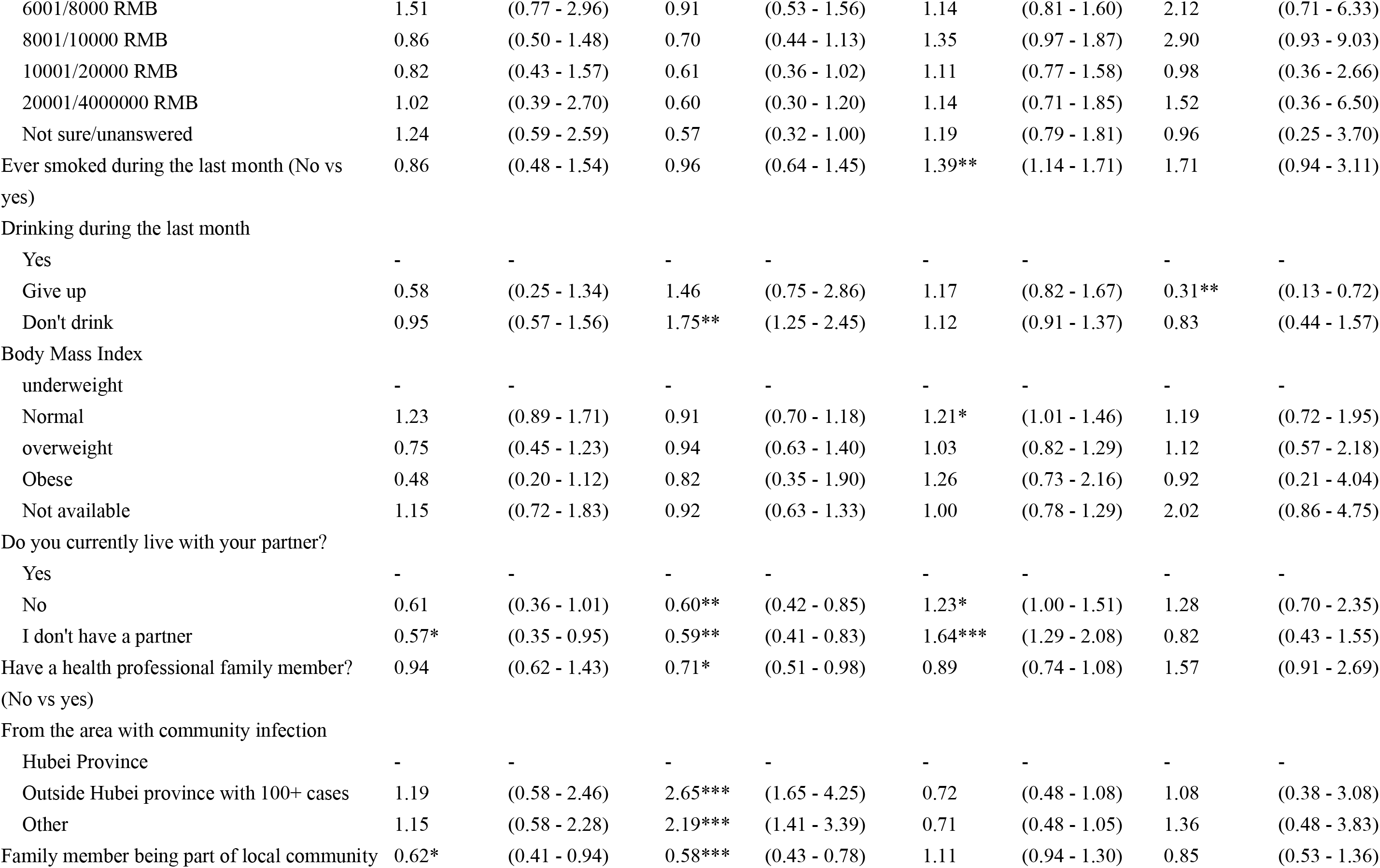
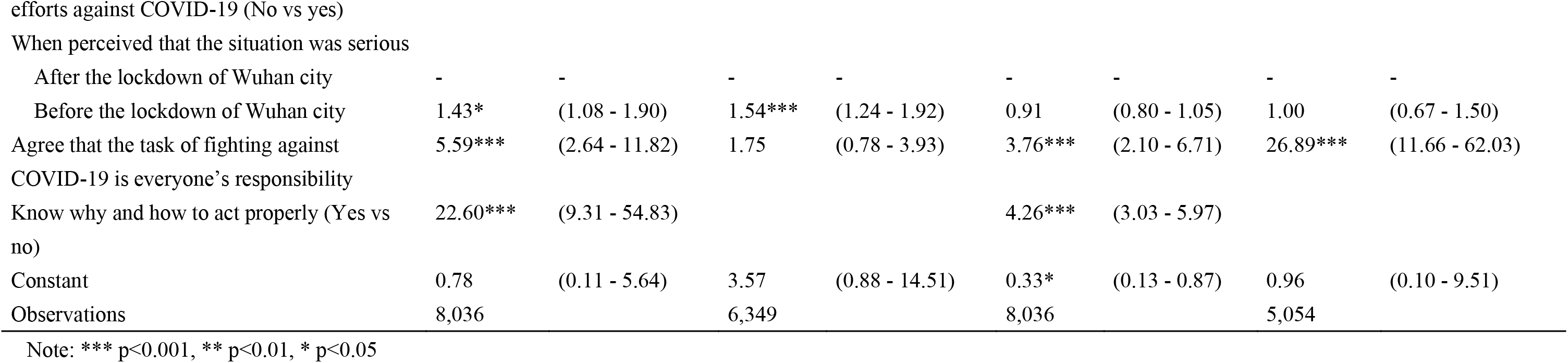
ORs and their 95%CI from logistic regression models for four NPIs.

| Predictors | Hands washing(yes/no) |  | Proper coughing(yes/no) |  | Social distancing(yes/no) |  | Mask Wearing(yes/no) |  |
| --- | --- | --- | --- | --- | --- | --- | --- | --- |
|  | OR | 95%CI | OR | 95%CI | OR | 95%CI | OR | 95%CI |
| Age groups |  |  |  |  |  |  |  |  |
| 18-39 years | - | - | - | - | - | - | - | - |
| 40-59 years | 1.46 | (0.84 - 2.53) | 2.24*** | (1.56 - 3.23) | 0.76** | (0.64 - 0.91) | 1.08 | (0.65 - 1.82) |
| >=60 years | 0.83 | (0.27 - 2.51) | 2.50 | (0.97 - 6.44) | 0.51*** | (0.35 - 0.73) | 0.25** | (0.10 - 0.63) |
| Sex (Female vs male) | 1.12 | (0.82 - 1.53) | 0.99 | (0.78 - 1.27) | 1.27** | (1.08 - 1.50) | 0.74 | (0.43 - 1.26) |
| Living in city (vs rural) | 1.67*** | (1.26 - 2.22) | 0.97 | (0.76 - 1.22) | 0.93 | (0.79 - 1.10) | 1.86** | (1.18 - 2.94) |
| Education |  |  |  |  |  |  |  |  |
| Primary School | - | - | - | - | - | - | - | - |
| High School | 2.22 | (0.75 - 6.53) | 2.72* | (1.20 - 6.16) | 1.08 | (0.67 - 1.76) | 0.59 | (0.10 - 3.32) |
| Professional college | 3.12 | (0.97 - 10.09) | 5.77*** | (2.39 - 13.91) | 1.30 | (0.79 - 2.16) | 0.46 | (0.08 - 2.73) |
| University/Postgraduate | 1.68 | (0.55 - 5.15) | 2.92* | (1.25 - 6.83) | 1.31 | (0.79 - 2.19) | 0.68 | (0.11 - 4.16) |
| Occupation |  |  |  |  |  |  |  |  |
| Health Professionals | - | - | - | - | - | - | - | - |
| Government payee | 0.41 | (0.15 - 1.15) | 0.78 | (0.44 - 1.38) | 0.85 | (0.67 - 1.08) | 1.32 | (0.72 - 2.43) |
| Factory workers/Managers | 0.23** | (0.08 - 0.66) | 0.59 | (0.33 - 1.08) | 0.81 | (0.61 - 1.06) | 1.22 | (0.57 - 2.60) |
| Farmers | 0.31 | (0.08 - 1.19) | 0.80 | (0.32 - 2.00) | 0.89 | (0.59 - 1.33) | 0.75 | (0.24 - 2.38) |
| Students | 0.10*** | (0.03 - 0.28) | 0.30*** | (0.16 - 0.55) | 0.61** | (0.44 - 0.86) | 1.24 | (0.48 - 3.19) |
| Others | 0.28* | (0.09 - 0.86) | 0.71 | (0.37 - 1.37) | 1.00 | (0.73 - 1.35) | 1.24 | (0.50 - 3.11) |
| Family monthly Income |  |  |  |  |  |  |  |  |
| 0/1000 RMB | - | - | - | - | - | - | - | - |
| 1001/2000 RMB | 1.45 | (0.82 - 2.56) | 0.74 | (0.47 - 1.19) | 0.91 | (0.68 - 1.23) | 1.06 | (0.46 - 2.47) |
| 2001/4000 RMB | 1.13 | (0.70 - 1.81) | 0.72 | (0.47 - 1.09) | 1.01 | (0.77 - 1.33) | 1.03 | (0.48 - 2.22) |
| 4001/6000 RMB | 0.84 | (0.52 - 1.34) | 0.89 | (0.57 - 1.37) | 1.01 | (0.76 - 1.35) | 1.19 | (0.53 - 2.69) |
| 6001/8000 RMB | 1.51 | (0.77 - 2.96) | 0.91 | (0.53 - 1.56) | 1.14 | (0.81 - 1.60) | 2.12 | (0.71 - 6.33) |
| 8001/10000 RMB | 0.86 | (0.50 - 1.48) | 0.70 | (0.44 - 1.13) | 1.35 | (0.97 - 1.87) | 2.90 | (0.93 - 9.03) |
| 10001/20000 RMB | 0.82 | (0.43 - 1.57) | 0.61 | (0.36 - 1.02) | 1.11 | (0.77 - 1.58) | 0.98 | (0.36 - 2.66) |
| 20001/4000000 RMB | 1.02 | (0.39 - 2.70) | 0.60 | (0.30 - 1.20) | 1.14 | (0.71 - 1.85) | 1.52 | (0.36 - 6.50) |
| Not sure/unanswered | 1.24 | (0.59 - 2.59) | 0.57 | (0.32 - 1.00) | 1.19 | (0.79 - 1.81) | 0.96 | (0.25 - 3.70) |
| Ever smoked during the last month (No vs yes) | 0.86 | (0.48 - 1.54) | 0.96 | (0.64 - 1.45) | 1.39** | (1.14 - 1.71) | 1.71 | (0.94 - 3.11) |
| Drinking during the last month |  |  |  |  |  |  |  |  |
| Yes | - | - | - | - | - | - | - | - |
| Give up | 0.58 | (0.25 - 1.34) | 1.46 | (0.75 - 2.86) | 1.17 | (0.82 - 1.67) | 0.31** | (0.13 - 0.72) |
| Don't drink | 0.95 | (0.57 - 1.56) | 1.75** | (1.25 - 2.45) | 1.12 | (0.91 - 1.37) | 0.83 | (0.44 - 1.57) |
| Body Mass Index |  |  |  |  |  |  |  |  |
| underweight | - | - | - | - | - | - | - | - |
| Normal | 1.23 | (0.89 - 1.71) | 0.91 | (0.70 - 1.18) | 1.21* | (1.01 - 1.46) | 1.19 | (0.72 - 1.95) |
| overweight | 0.75 | (0.45 - 1.23) | 0.94 | (0.63 - 1.40) | 1.03 | (0.82 - 1.29) | 1.12 | (0.57 - 2.18) |
| Obese | 0.48 | (0.20 - 1.12) | 0.82 | (0.35 - 1.90) | 1.26 | (0.73 - 2.16) | 0.92 | (0.21 - 4.04) |
| Not available | 1.15 | (0.72 - 1.83) | 0.92 | (0.63 - 1.33) | 1.00 | (0.78 - 1.29) | 2.02 | (0.86 - 4.75) |
| Do you currently live with your partner? |  |  |  |  |  |  |  |  |
| Yes | - | - | - | - | - | - | - | - |
| No | 0.61 | (0.36 - 1.01) | 0.60** | (0.42 - 0.85) | 1.23* | (1.00 - 1.51) | 1.28 | (0.70 - 2.35) |
| I don't have a partner | 0.57* | (0.35 - 0.95) | 0.59** | (0.41 - 0.83) | 1.64*** | (1.29 - 2.08) | 0.82 | (0.43 - 1.55) |
| Have a health professional family member? (No vs yes) | 0.94 | (0.62 - 1.43) | 0.71* | (0.51 - 0.98) | 0.89 | (0.74 - 1.08) | 1.57 | (0.91 - 2.69) |
| From the area with community infection |  |  |  |  |  |  |  |  |
| Hubei Province | - | - | - | - | - | - | - | - |
| Outside Hubei province with 100+ cases | 1.19 | (0.58 - 2.46) | 2.65*** | (1.65 - 4.25) | 0.72 | (0.48 - 1.08) | 1.08 | (0.38 - 3.08) |
| Other | 1.15 | (0.58 - 2.28) | 2.19*** | (1.41 - 3.39) | 0.71 | (0.48 - 1.05) | 1.36 | (0.48 - 3.83) |
| Family member being part of local community | 0.62* | (0.41 - 0.94) | 0.58*** | (0.43 - 0.78) | 1.11 | (0.94 - 1.30) | 0.85 | (0.53 - 1.36) |
| After the lockdown of Wuhan city | - | - | - | - | - | - | - | - |
| Before the lockdown of Wuhan city | 1.43* | (1.08 - 1.90) | 1.54*** | (1.24 - 1.92) | 0.91 | (0.80 - 1.05) | 1.00 | (0.67 - 1.50) |
| Agree that the task of fighting against COVID-19 is everyone's responsibility | 5.59*** | (2.64 - 11.82) | 1.75 | (0.78 - 3.93) | 3.76*** | (2.10 - 6.71) | 26.89*** | (11.66 - 62.03) |
| Know why and how to act properly (Yes vs no) | 22.60*** | (9.31 - 54.83) |  |  | 4.26*** | (3.03 - 5.97) |  |  |
| Constant | 0.78 | (0.11 - 5.64) | 3.57 | (0.88 - 14.51) | 0.33* | (0.13 - 0.87) | 0.96 | (0.10 - 9.51) |
| Observations | 8,036 |  | 6,349 |  | 8,036 |  | 5,054 |  |

Note: \*\*\* p<0.001, \*\* p<0.01, \* p<0.05

**eTable 2.** Self-isolation.

| Questions: | N=8158 | % |
| --- | --- | --- |
| Had party during new year eve and new year (2 days) |  |  |
| Yes | 1833 | 22.5% |
| No | 6242 | 76.5% |
| Not sure | 83 | 1.0% |
| The major reason for staying at home |  |  |
| self/compulsory insolation | 1107 | 13.6% |
| Fear of the virus | 469 | 5.7% |
| The focus of family protection | 310 | 3.8% |
| No mask | 242 | 3.0% |
| Ill health | 63 | 0.8% |
| Comply with the call from government | 5354 | 65.6% |
| Other | 613 | 7.5% |
| The main reason for went out* |  |  |
| Shopping | 2073 | 40.5% |
| Partaking the job against COVID-19 | 1643 | 32.1% |
| Doing usual job | 1098 | 21.4% |
| Getting delivery | 38 | 0.7% |
| Socializing/Dinner party | 17 | 0.3% |
| Having a walk | 131 | 2.6% |
| Other | 120 | 2.3% |
| What did you do when you went out* |  |  |
| Shorten the time to avoid infection | 3798 | 74.2% |
| As usual | 1000 | 19.5% |
| Stay longer given the hassle | 50 | 1.0% |
| Uncertain | 272 | 5.3% |

\*Only including 5120 respondents who went out after the outbreak of COVID-19.

**eTable 3.**
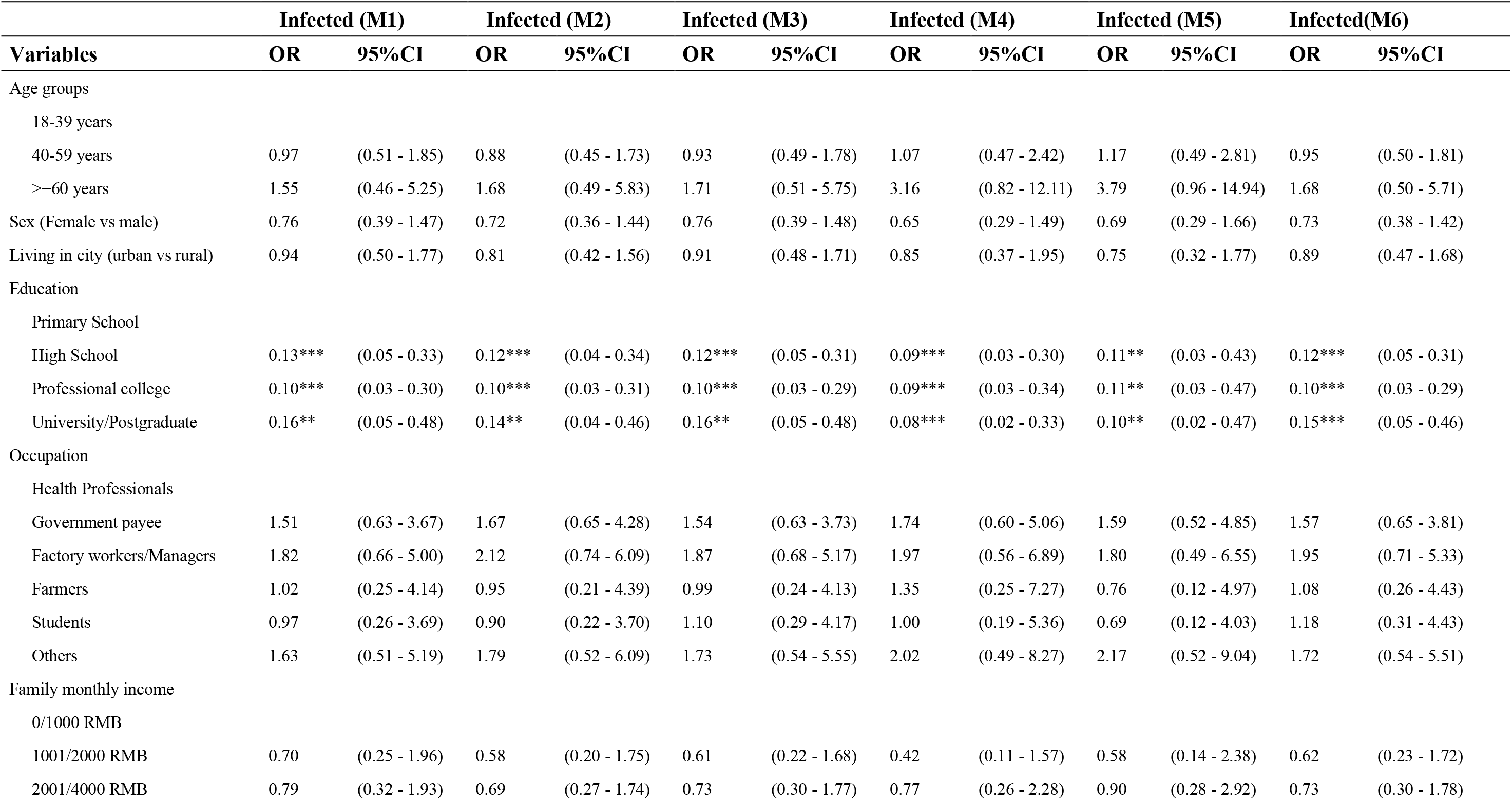
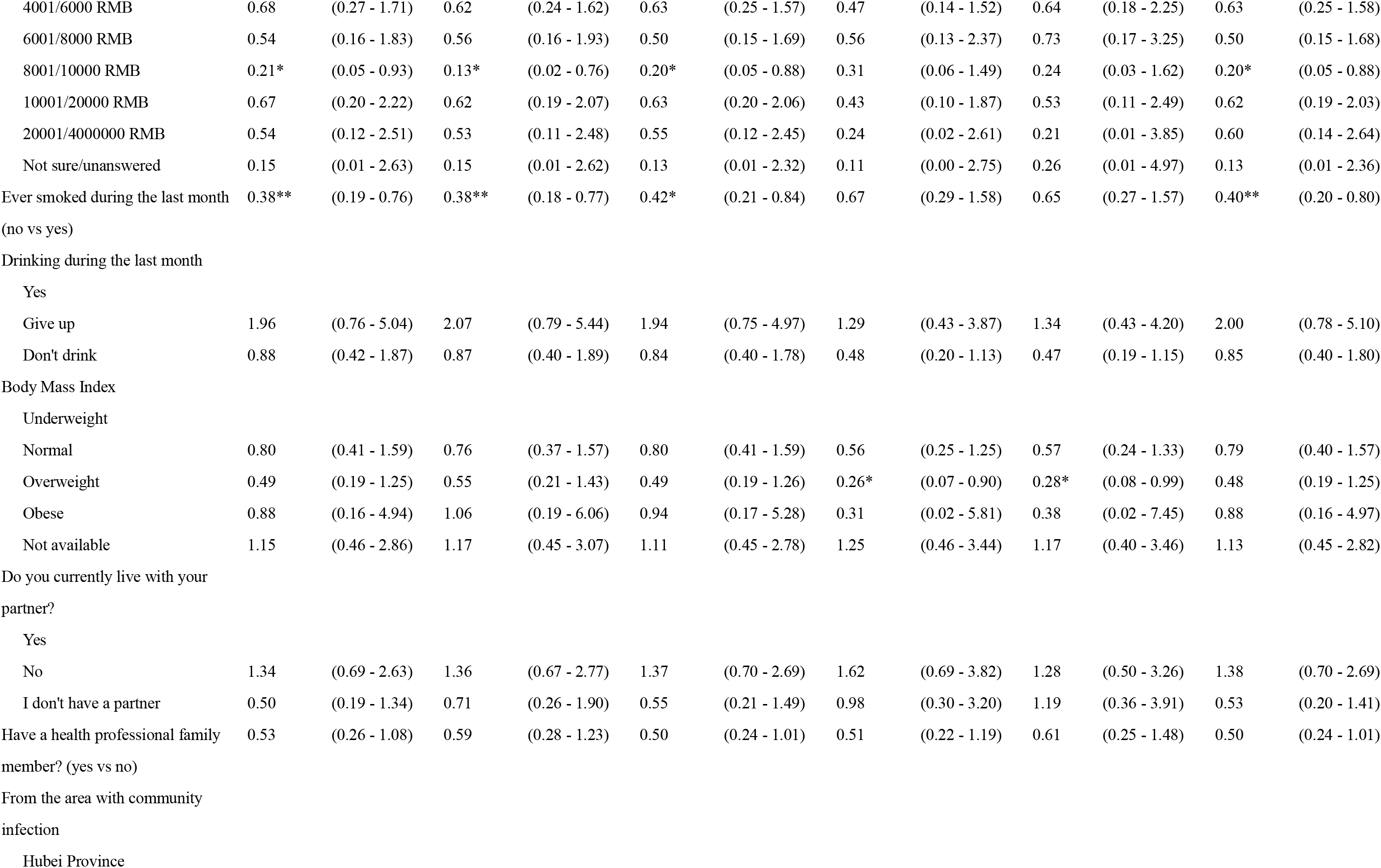
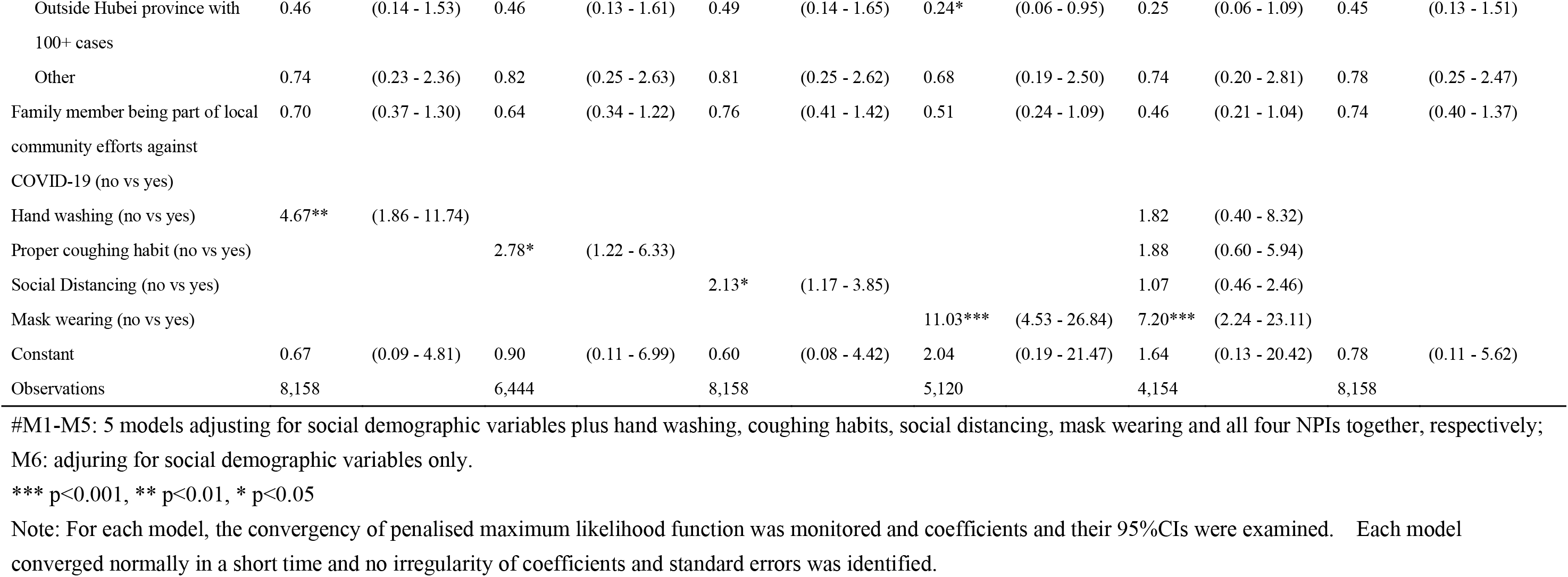
Penalised logistic regression model results for COVID-19 infection and hand washing, coughing habits, social distancing, mask wearing^#^.

